# Integrative Multimodal Subtyping, Risk of Incident Mild Cognitive Impairment, and Differential Cardiometabolic Treatment Effects: A Prospective Cohort Study in the All of Us Research Program

**DOI:** 10.64898/2026.02.10.26345240

**Authors:** Yinjun Zhao, Karen Marder, Yuanjia Wang

## Abstract

**Background:** Cognitively unimpaired (CU) adults vary substantially in their risk of developing mild cognitive impairment (MCI), yet most subtyping approaches focus on downstream neurobiological or cognitive markers rather than upstream, modifiable risk factors. We aimed to identify clinically meaningful subgroups of CU adults defined by integrated comorbid, behavioral, and social risk profiles, and to evaluate heterogeneity in both incident MCI risk and cardiometabolic treatment effects.

**Methods:** We conducted a prospective cohort study of 121,322 CU adults aged ≥50 years from the All of Us Research Program. Baseline comorbidities, lifestyle behaviors, and social determinants of health were jointly modeled using the Bayesian Mixed Integrative Data Subtyping framework, which integrates binary and continuous modalities via modality-specific likelihoods and shared latent constructs. Subtype-specific risk of incident MCI was assessed using multivariable Cox proportional hazards models adjusting for demographics and baseline medication use. A double/debiased machine learning interactive regression model with inverse probability of censoring weights to mitigate bias from informative censoring was implemented to estimate the average treatment effects of antihypertensive agents, Glucagon-Like Peptide (GLP) receptor agonists, and non–GLP antidiabetic medications on time to MCI.

**Results:** Four distinct subtypes were identified: I low-risk healthy aging, II behavioral/social vulnerability, III cardiometabolic–depressive multimorbidity, and IV mixed social-medical vulnerability profiles. Compared with Subtype I, Subtype III demonstrated the highest risk of incident MCI (HR: 3.69, 95% CI: 3.14–4.33), followed by Subtype IV and Subtype II. In treatment effect analyses, antihypertensive use was associated with a modest prolongation of MCI-free survival overall (time ratio:1.04, 95% CI: 1.03–1.06), with the largest benefit observed in Subtype III (time ratio: 1.14, 95% CI: 1.09–1.19). Non–GLP antidiabetic therapies were similarly associated with modest overall delay, with significant benefits in Subtypes I and III. GLP-class therapies were not associated with overall delay but showed a significant association in Subtype III.

**Conclusions:** Integrative subtyping based on comorbid, behavioral, and social risk factors reveals clinically meaningful heterogeneity in both cognitive risk and treatment response. Aligning dementia prevention strategies with dominant vulnerability pathways may enhance the effectiveness and equity of population-level precision prevention.

## Background

Mild cognitive impairment (MCI) is a common intermediate stage between normal aging and dementia, affecting approximately 15–20% of U.S. adults aged 60 years or older(1). Because disease-modifying therapies for dementia remain limited, with only two monoclonal antibody treatments currently approved and in clinical use after the withdrawal of aducanumab from the market(2), prevention efforts increasingly focus on identifying cognitively unimpaired (CU) individuals at elevated risk for future cognitive decline. In this context, accurate risk stratification among CU adults is central to targeted prevention, trial enrichment, and efficient allocation of preventive resources. Cognitive impairment and dementia are influenced by a broad range of modifiable risk factors, including cardiometabolic disease, depression, sensory impairment, social isolation, and socioeconomic disadvantage(2,3). A growing body of data-driven research has demonstrated substantial heterogeneity among CU individuals with respect to subsequent risk of MCI and dementia. Most existing subtyping studies in CU populations have concentrated on neurobiological and cognitive modalities, including structural magnetic resonance imaging, such as cortical thickness and regional brain volume; amyloid and tau positron emission tomography; fluid biomarkers, including cerebrospinal fluid and blood-based biomarkers; and detailed neuropsychological profiles(4–7). Using unsupervised clustering, latent class models, and disease-progression frameworks, these studies have shown that individuals who are clinically normal at baseline may nonetheless occupy distinct biomarker- or cognition-defined subgroups with markedly different longitudinal trajectories and rates of conversion. However, by focusing on downstream pathology, these approaches offer limited insight into upstream, modifiable drivers of risk and have restricted utility for population-level prevention.

In contrast, epidemiologic research has consistently implicated comorbid disease burden, lifestyle behaviors, and social determinants of health (SDOH) as central contributors to cognitive aging. Multimorbidity, particularly cardiometabolic and vascular conditions, has been robustly associated with elevated risk of incident MCI and dementia in large population-based cohorts(8–10). Lifestyle factors such as smoking, physical inactivity, and obesity further accelerate cognitive decline and account for a substantial proportion of potentially preventable dementia cases(11). Socioeconomic disadvantage, neighborhood deprivation, and psychosocial stress have likewise been linked to increased dementia incidence and faster cognitive aging(2,12). Despite this strong evidence base, most studies treat these factors as covariates or examine them in isolation, emphasizing population-average associations and obscuring the co-occurring vulnerability profiles and distinct risk pathways present in real-world populations(2,13,14).

The All of Us Research Program provides a unique opportunity to address this gap. As a national precision-medicine initiative designed to reflect the diversity of the U.S. population, All of Us integrates electronic health records with detailed lifestyle and SDOH survey data, enabling large-scale evaluation of how biological, behavioral, and social factors jointly shape cognitive risk(15). However, these data are high-dimensional, correlated, and measured on mixed scales, posing challenges for conventional clustering and feature-based multimodal methods that are not designed to jointly model heterogeneous data types. To address these challenges, we apply a Bayesian integrative subtyping framework (MINDS) that jointly models binary and continuous variables and identifies latent constructs capturing shared variation across multimodal domains(16). By integrating comorbidity burden, lifestyle behaviors, and SDOH indicators into a unified latent space, MINDS enables discovery of interpretable subtypes that reflect real-world vulnerability profiles rather than single-modality patterns. In this study, we sought to (1) identify clinically meaningful subtypes among cognitively unimpaired adults using integrated modeling of comorbidities, lifestyle factors, and SDOH; (2) evaluate heterogeneity in incident MCI risk across these subtypes; and (3) assess whether commonly used cardiometabolic therapies antihypertensive, GLP-1 receptor agonist, and non–GLP antidiabetic medications demonstrate differential associations with time to MCI across vulnerability profiles. We focused on these medications because hypertension and type 2 diabetes are established upstream cardiometabolic risk factors for cognitive decline and dementia(2,13,17,18). These therapies have shown potential to delay cognitive impairment in both randomized trials and large meta-analyses(14,19–21). By explicitly modeling heterogeneity in both baseline risk and treatment responsiveness, this work advances a population-level framework for precision dementia prevention and contributes to understanding how upstream, modifiable risk factors jointly shape cognitive aging.

## Methods

### Integrative subtyping framework: MINDS

To identify data-driven subtypes that capture heterogeneity across comorbidities, lifestyle behaviors, and social determinants of health, we employed the Mixed INtegrative Data Subtyping (MINDS) framework, a Bayesian hierarchical model for joint analysis of mixed-type data(16). MINDS was originally developed to integrate multimodal clinical, cognitive, and neuroimaging data for psychiatric subtyping and is designed to perform simultaneous dimension reduction and clustering while preserving shared structure across heterogeneous modalities.

At its core, MINDS models each data modality through a modality-specific likelihood, i.e., Bernoulli for binary indicators and Gaussian for continuous measures, linked by shared cluster-specific latent centers and subject-specific latent construct. Individuals are assumed to belong to one of the latent clusters, each characterized by distinct mean profiles in the latent construct space, while allowing subject-specific deviations. This formulation enables coherent probabilistic integration of diverse data types without requiring ad hoc transformations or two-stage clustering procedures. Computationally, MINDS leverages Pólya–Gamma data augmentation to yield conditionally conjugate posterior distributions for binary outcomes, enabling efficient Gibbs sampling for posterior inference(22). The number of subtypes is selected using a Bayesian information criterion (IC) that balances model fit and parsimony, as described in the original methodological work(16).

### Cohort for subtyping

We applied MINDS to identify data-driven subtypes among participants in the All*of*Us Research Program by jointly integrating comorbidities, SDOH, and lifestyle factors. The Curated Data Repository Version 8, released by the All*of*Us Research Program and containing participant data through October 1, 2023, served as the source for all analyses(23). We began with all eligible participants aged 50 years or older and linked their electronic health records (EHR), SDOH survey responses, and lifestyle survey data. Participants missing an entire modality at baseline or with documented cognitive function finding at baseline were excluded to reduce potential bias arising from systematically different health or survey-response profiles. Participants with any recorded cognitive function finding were defined as the SNOMED CT parent concept “Cognitive function finding” (OMOP concept id 4162723) and all its descendant concepts, identified in the OMOP CONDITION domain. This definition captures clinician-documented cognitive abnormalities (e.g., memory, attention, or thought findings). The final analytic cohort consisted of 121,322 individuals with complete multimodal data and follow-ups to assess incident MCI. Figure 1 depicts the derivation of this cohort.

**Figure 1:** Flowchart of participant selection and exclusion criteria in the All of Us cohort.

Comorbidity status at baseline including stroke, type 2 diabetes, heart failure, hearing loss, depression, hypertension, and brain injury was obtained from EHR data. Lifestyle behaviors were extracted from the lifestyle survey. Among current drinkers, we constructed the AUDIT–C score (range, 0–12) using three items assessing (1) drinking frequency, (2) typical quantity consumed per drinking occasion, and (3) frequency of binge drinking (defined as consuming 6 or more drinks on a single occasion). Each item included five ordered response options scored from 0 (lower consumption) to 4 (higher consumption). Alcohol misuse was defined as an AUDIT–C score of 3 or higher for women and 4 or higher for men(24). Current smoker status was obtained from the corresponding lifestyle survey question. SDOH measures were obtained from validated instruments included in the SDOH survey. These measures encompassed four conceptual domains: social and community context, economic stability, neighborhood and built environment, and health and healthcare(25). Constructs such as social cohesion, social support, loneliness, perceived discrimination, perceived stress, and daily spiritual experiences were assessed through established multi-item scales. Economic stability was evaluated using indicators of food insecurity, housing instability, and housing quality. Neighborhood characteristics, including physical and social disorder, walk ability, and crime, were captured using standardized environmental assessment tools, and perceived discrimination in healthcare settings was assessed using a dedicated subscale. Higher values of the SDOH measures reflected worse situation, consistent with the direction of comorbidity and lifestyle measures. Detailed descriptions of each construct and measurement instrument are provided in **Error! Reference source not found.**. Collectively, these multidimensional measures served as inputs for the subtyping analysis.

### MCI association analysis

To investigate the prognostic relevance of the identified subtypes, we conducted an exploratory survival analysis to examine the association between subtypes and the subsequent risk of developing MCI. Time-to-event was defined as the interval from cohort entry to the first documented diagnosis of MCI or censoring, whichever occurred earlier. Kaplan–Meier curves were generated to provide an initial descriptive comparison of incident MCI across clusters. We then fit Cox proportional hazards models to estimate hazard ratios (HRs) and 95% confidence intervals (CIs) for MCI associated with each subtype, controlling for demographics and antihypertensive, non-GLP antidiabetic and GLP class medication use at baseline. The medication use was ascertained from electronic health records based on prescriptions recorded within one year before the baseline assessment. Participants with evidence of use of any medication in a given class during this one-year window before baseline were classified as users of that medication class. Details of the agents that were included in the medication class are in **Error! Reference source not found.**.

### Treatment effect analysis

We estimated the causal effects of antihypertensives, non–GLP antidiabetic, and GLP-class medications use on time to incident MCI using a double/debiased machine learning (DoubleML) interactive regression model (IRM)(26). DoubleML is a causal inference approach designed for observational studies with complex, high-dimensional confounding. The method leverages flexible machine-learning algorithms to model both treatment assignment and outcome processes (“double”) and combines them in a way that reduces bias from model misspecification (“debiased”). It ensures valid causal inference when either the treatment model or the outcome model is correctly specified, increasing robustness in observational data analyses. Medication use was treated as a binary exposure, and confounding variables included age, sex, race/ethnicity, educational attainment, and baseline cardiometabolic comorbidities.

The primary estimand was the average treatment effect (ATE), defined as the expected difference in outcomes had all individuals received versus not received the treatment. To assess heterogeneity in treatment response, we additionally estimated group average treatment effects (GATEs), which quantify treatment effects within each data-driven subtype identified by the integrative subtyping model. The outcome was defined as the logarithm of observed follow-up time to MCI, such that treatment effects can be interpreted on a multiplicative time scale. Exponentiating the estimated effects yields a geometric mean time ratio, representing the relative prolongation (or shortening) of MCI-free survival associated with medication use.

To account for right censoring, we incorporated inverse probability of censoring weights (IPCW). Specifically, we estimated the probability of remaining uncensored over time conditional on treatment status and covariates, and weighted complete observations by the inverse of this probability. This procedure reduces bias due to informative censoring under standard assumptions(27). A fixed cutoff time of 48 months was used to define the at-risk set, including individuals who either experienced the event before this time or remained under observation beyond it.

## Results

### Subtyping results

We evaluated models with the number of clusters ranging from *k* == 3 to *k* == 9 using IC(28) to determine the optimal cluster structure. As shown in Supple. **Error! Reference source not found.**(a), the IC reached its minimum at *k* == 4, indicating that the four-cluster model achieved the best balance between model fit and parsimony. To assess convergence and model stability, we examined posterior log-likelihood trace plot in Supple. **Error! Reference source not found.**(b), which demonstrated stable convergence and consistent mixing of the Markov chain Monte Carlo (MCMC) samples after the burn-in period. These results suggest that the four-cluster solution provides a well-fitted and robust model for integrating the multimodal data.

As shown in Figure 2, the four subtypes demonstrated distinct and coherent patterns across comorbidity burden, lifestyle risk behaviors, and social determinants of health. Subtype I reflected a broadly healthy profile, characterized by minimal chronic disease, low rates of smoking and alcohol misuse, and consistently favorable social and environmental conditions, suggesting a Low-Risk Healthy Aging group. Subtype II showed markedly elevated smoking and alcohol misuse alongside the highest levels of social and economic adversity, including neighborhood disorder, discrimination, social isolation, and housing instability, indicating a Behavioral-Risk and Socially Vulnerable subgroup. Subtype III exhibited the greatest medical burden, with pronounced hypertension, depression, hearing loss, and cardiometabolic disease, paired with moderate psychosocial stressors, aligning with a Cardiometabolic–Depressive Multimorbidity subtype. Finally, Subtype IV demonstrated intermediate levels of comorbidity and moderately elevated SDOH disadvantage, representing a Mixed-Risk Mild Vulnerability group. Together, these patterns highlight the multidimensional heterogeneity in health, lifestyle, and social context within the population. The characteristics of the four types are summarized in Table 3, which are also reflected in the summary statistics by the four subtypes in Supple. **Error! Reference source not found.**.

**Figure 2:** Heatmaps of binary and continuous measures across subtypes, integrating lifestyle factors, comorbidity and SDOH.

The radar plots provide a visual comparison of subtype differences across SDOH, lifestyle, and comorbidity measures in another way. The first plot (Figure 3) illustrates variation in social determinants of health across the four subtypes, highlighting pronounced contrasts in constructs such as discrimination in healthcare, social support, social cohesion, perceived stress, loneliness, neighborhood disorder, and material hardship (e.g., food insecurity, housing instability). Subtypes II and IV show elevated levels on multiple adverse SDOH indicators, whereas Subtype I generally reflects more favorable social and environmental conditions. The second plot (Figure 3) summarizes subtype differences in lifestyle risk factors and chronic disease burden, including smoking, alcohol misuse, and major comorbidities such as hypertension, diabetes, heart failure, stroke, and depression. Subtype III shows elevated levels on comorbidities. Together, these plots highlight distinct multidimensional profiles across subtypes, demonstrating how combinations of social context, behavior, and health conditions jointly characterize the heterogeneity captured by the clustering model.

**Figure 3:** Radar plot illustrating differences in SDOH and comorbidity profiles across subtypes I–IV.

The three latent constructs that MINDS estimated reflect distinct but complementary domains of vulnerability. As shown in Figure 4, LC1 is best characterized as Neighborhood Disorder and Safety Risk, defined by high levels of neighborhood physical and social disorder and low perceptions of crime safety, indicating exposure to adverse environmental conditions. LC2 represents a pattern of High-Burden Chronic Multimorbidity, marked by strong associations with stroke, type 2 diabetes, heart failure, hypertension, hearing loss, brain injury, and depression. LC3 reflects Psychosocial Stress and Social Vulnerability, driven by elevated food insecurity, poor housing quality, loneliness, discrimination (every day and within health-care settings), perceived stress, and low social support and cohesion. See Supple. **Error! Reference source not found.** for the exact values of the loading matrix.

**Figure 4:** Heatmap displaying standardized loadings for each observed measure across the three latent constructs (LC1–LC3).

Supple. Figure A.2 illustrates the distribution of the three latent constructs (LC1–LC3) across the four subtypes, highlighting clear and interpretable subtype-specific patterns. Subtype I shows consistently low to moderate expression of LC1 and minimal contributions from LC2 and LC3, reflecting a comparatively low burden of adverse social and health factors. Subtype II demonstrates a stronger LC1 profile, driven by elevated social and psychological stressors such as loneliness, discrimination, and perceived stress. In contrast, Subtype III is distinguished by a dominant LC2 pattern, with high scores on multiple comorbidities including hypertension, type 2 diabetes, hearing loss, depression, and other chronic conditions. Subtype IV exhibits the strongest expression of LC3, characterized by markedly higher levels of neighborhood physical/social disorder and crime safety concerns. Together, these distinct profiles demonstrate that the subtypes are differentiated not only in magnitude but also in the composition of their latent constructs, capturing qualitatively different combinations of social, environmental, and health-related vulnerabilities.

### Subtype’s association with MCI

The models were first examined unadjusted (Supple. **Error! Reference source not found.**) and then adjusted for relevant demographic, clinical, and medication-use covariates(Table 2) to explore whether the observed associations persisted after accounting for potential confounders.

**Table 1:** Characteristics of the four data-driven subtypes based on comorbidity, lifestyle behaviors, and social determinants of health.

**Table 2:** Cox Regression Results on time to MCI by subtype, demographics, and medication use.

**Table 3:**
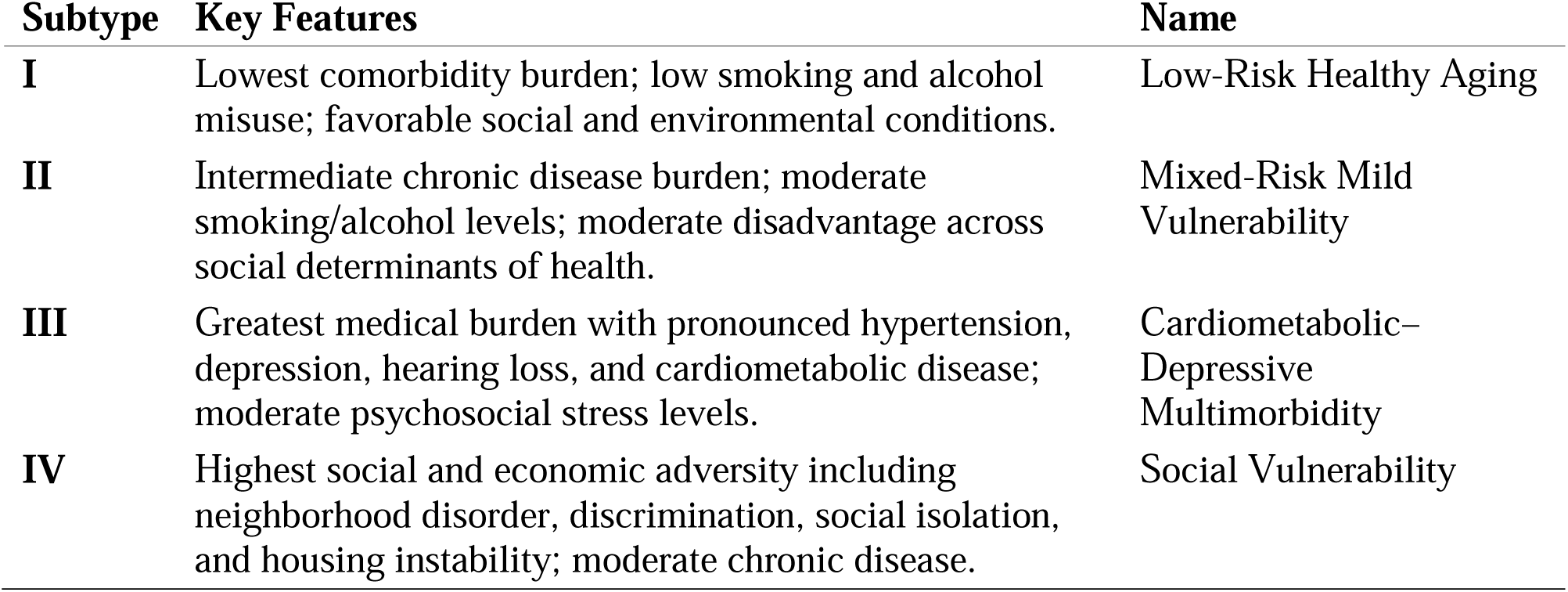
Characteristics of the four data-driven subtypes based on comorbidity, lifestyle behaviors, and social determinants of health.

| Subtype | Key Features | Name |
| --- | --- | --- |
| I | Lowest comorbidity burden; low smoking and alcohol misuse; favorable social and environmental conditions. | Low-Risk Healthy Aging |
| II | Intermediate chronic disease burden; moderate smoking/alcohol levels; moderate disadvantage across social determinants of health. | Mixed-Risk Mild Vulnerability |
| III | Greatest medical burden with pronounced hypertension, depression, hearing loss, and cardiometabolic disease; moderate psychosocial stress levels. | Cardiometabolic–Depressive Multimorbidity |
| IV | Highest social and economic adversity including neighborhood disorder, discrimination, social isolation, and housing instability; moderate chronic disease. | Social Vulnerability |

**Table 4:** Cox Regression Results on time to MCI by subtype, demographics, and medication use.

| Characteristic | N | HR | 95% CI | p-value |
| --- | --- | --- | --- | --- |
| **Subtype** |  |  |  | **<0.001** |
| I | 66,422 | — | — |  |
| II | 9,039 | 1.11 | 0.53, 2.32 |  |
| III | 30,070 | 4.71 | 3.40, 6.52 |  |
| IV | 15,791 | 1.96 | 1.17, 3.30 |  |
| **Age (years)** | 121,322 | 1.85 | 1.69, 2.03 | **<0.001** |
| **Gender** |  |  |  | **<0.001** |
| Female | 74,270 | — | — |  |
| Male | 47,052 | 1.37 | 1.16, 1.61 |  |
| **Race** |  |  |  | 0.9 |
| Non-White | 15,914 | — | — |  |
| White | 105,408 | 1.02 | 0.77, 1.35 |  |
| **Ethnicity** |  |  |  | 0.2 |
| Non-Hispanic | 110,172 | — | — |  |
| Hispanic | 11,150 | 1.22 | 0.88, 1.69 |  |
| **Educational attainment** |  |  |  | 0.5 |
| Below college | 46,794 | — | — |  |
| College & above | 74,528 | 0.94 | 0.79, 1.12 |  |
| **Antihypertensive use** |  |  |  | **<0.001** |
| No | 80,649 | — | — |  |
| Yes | 40,673 | 3.23 | 2.35, 4.42 |  |
| **Non-GLP antidiabetic use** |  |  |  | 0.5 |
| No | 108,002 | — | — |  |
| Yes | 13,320 | 1.13 | 0.77, 1.65 |  |
| **GLP-1 class use** |  |  |  | 0.077 |
| No | 116,665 | — | — |  |
| Yes | 4,657 | 2.16 | 1.01, 4.66 |  |
| **Subtype × Antihypertensive use** | 121,322 |  |  | **<0.001** |
| II * Yes | 2,605 | 1.25 | 0.51, 3.07 |  |
| III * Yes | 14,025 | 0.31 | 0.21, 0.46 |  |
| IV * Yes | 5,713 | 0.87 | 0.47, 1.62 |  |
| **Subtype × Non-GLP antidiabetic use** | 121,322 |  |  | >0.9 |
| II * Yes | 842 | 1.19 | 0.45, 3.10 |  |
| III * Yes | 4,576 | 0.94 | 0.57, 1.54 |  |
| IV * Yes | 2,071 | 1.13 | 0.58, 2.18 |  |
| **Subtype × GLP class use** | 121,322 |  |  | 0.7 |
| II * Yes | 239 | 0.44 | 0.05, 3.79 |  |
| III * Yes | 2,418 | 0.71 | 0.30, 1.67 |  |
| IV * Yes | 928 | 0.58 | 0.19, 1.72 |  |

The four data-driven subtypes demonstrated clear heterogeneity in their risk of developing MCI. Using Subtype I as the reference, individuals in Subtype III exhibited the highest risk, with HR 3.69 (95% CI: 3.14–4.33), indicating more than a threefold increase in the hazard of MCI. Subtype IV also showed significantly elevated risk (HR = 1.87, 95% CI: 1.48–2.35), whereas Subtype II demonstrated a more modest but still significant increase (HR = 1.43, 95% CI: 1.07–1.91). These subtype differences were further reflected in the Kaplan–Meier survival curves in Figure 5, where Subtype III displayed the steepest decline in MCI-free survival, followed by Subtype IV and Subtype II, while Subtype I maintained the most favorable survival trajectory. Together, these findings indicate that Subtype III represents a particularly vulnerable group with accelerated cognitive decline, whereas Subtype I appears comparatively resilient, with Subtypes II and IV occupying intermediate risk profiles.

**Figure 5:** Kaplan–Meier survival curves depicting the probability of remaining free from MCI across Subtypes I–IV.

### Causal treatment effect

Figure 6 displays the estimated exponential ATEs of antihypertensive, non–GLP antidiabetic, and GLP-class medications on time to incident MCI in the overall cohort and across four data-driven subtypes. Antihypertensive use led a modestly longer time to MCI overall (time ratio = 1.04; 95% CI, 1.03–1.06), corresponding to an approximately 4% increase in geometric-mean follow-up time(months). Subtype III showed a markedly stronger association (time ratio = 1.14; 95% CI, 1.09–1.19; ∼14% longer time), while Subtype IV demonstrated a smaller but statistically significant association (time ratio = 1.04; 95% CI, 1.00–1.08). No significant associations were observed for Subtypes I or II.

**Figure 6:** Estimated exponential average treatment effects for antihypertensive, non-GLP antidiabetic and GLP class medications across subtypes.

**Figure 7:**
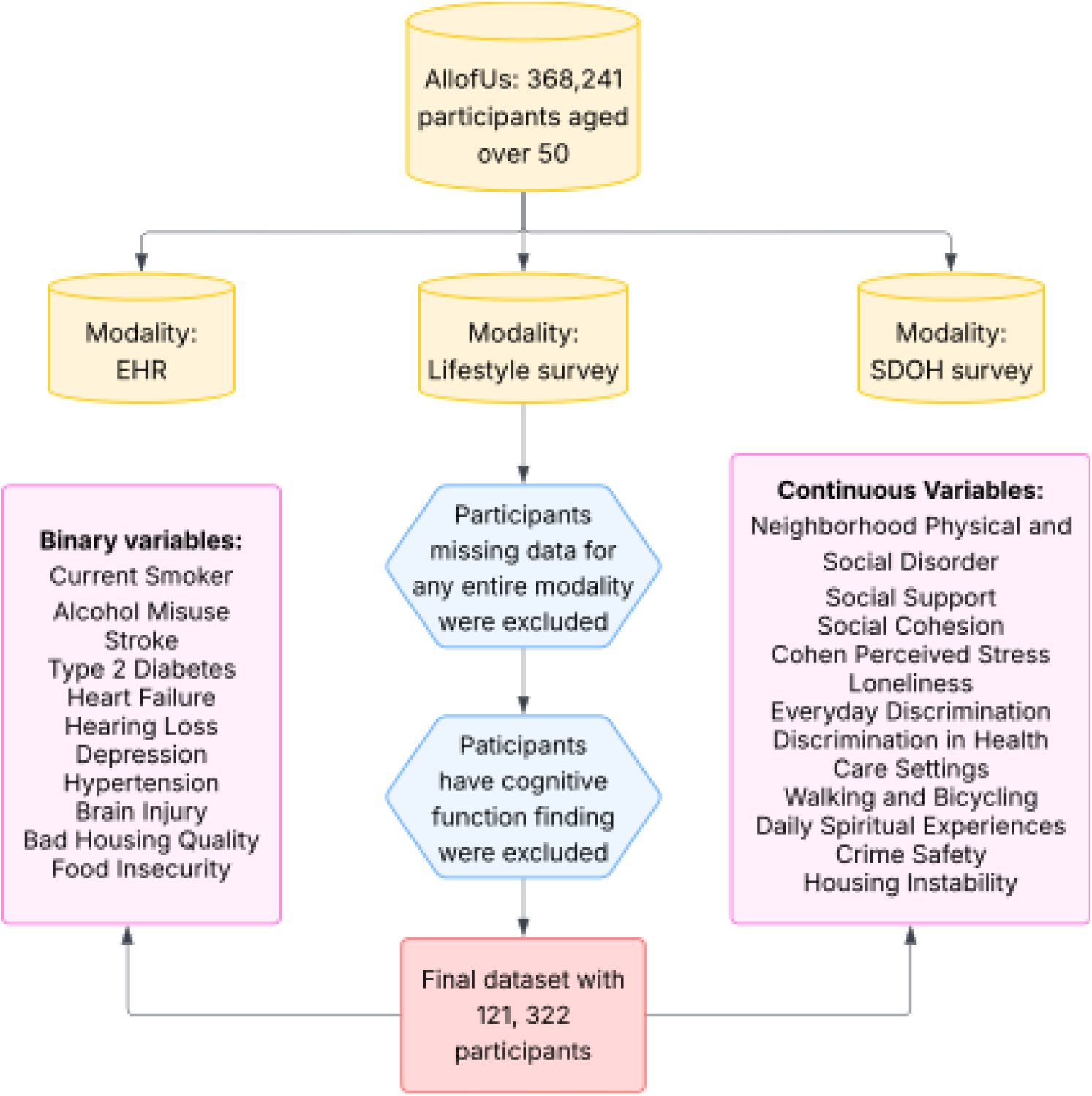
Flowchart of participant selection and exclusion criteria in the All of Us cohort.

**Figure 8:**
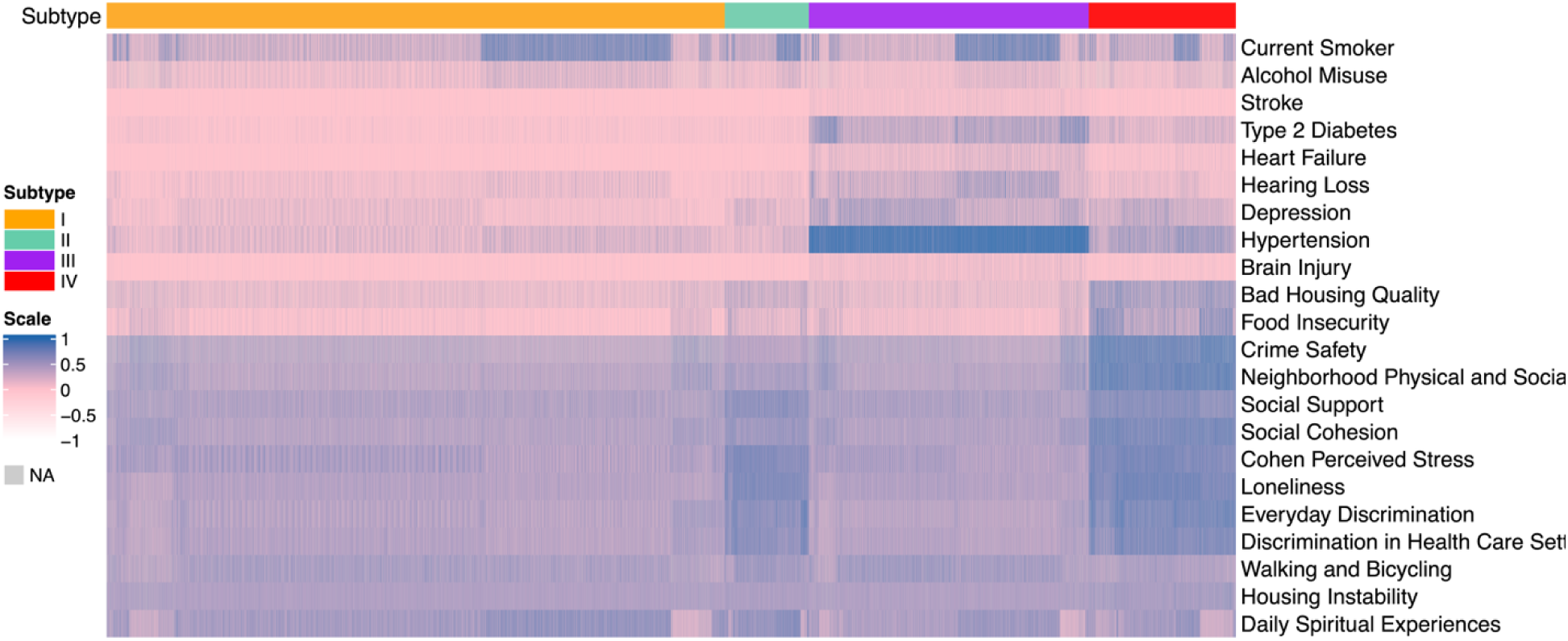
Heatmaps of binary and continuous measures across subtypes by the MINDS method, integrating lifestyle factors, comorbidity and social determinants of health measures.

**Figure 9:**
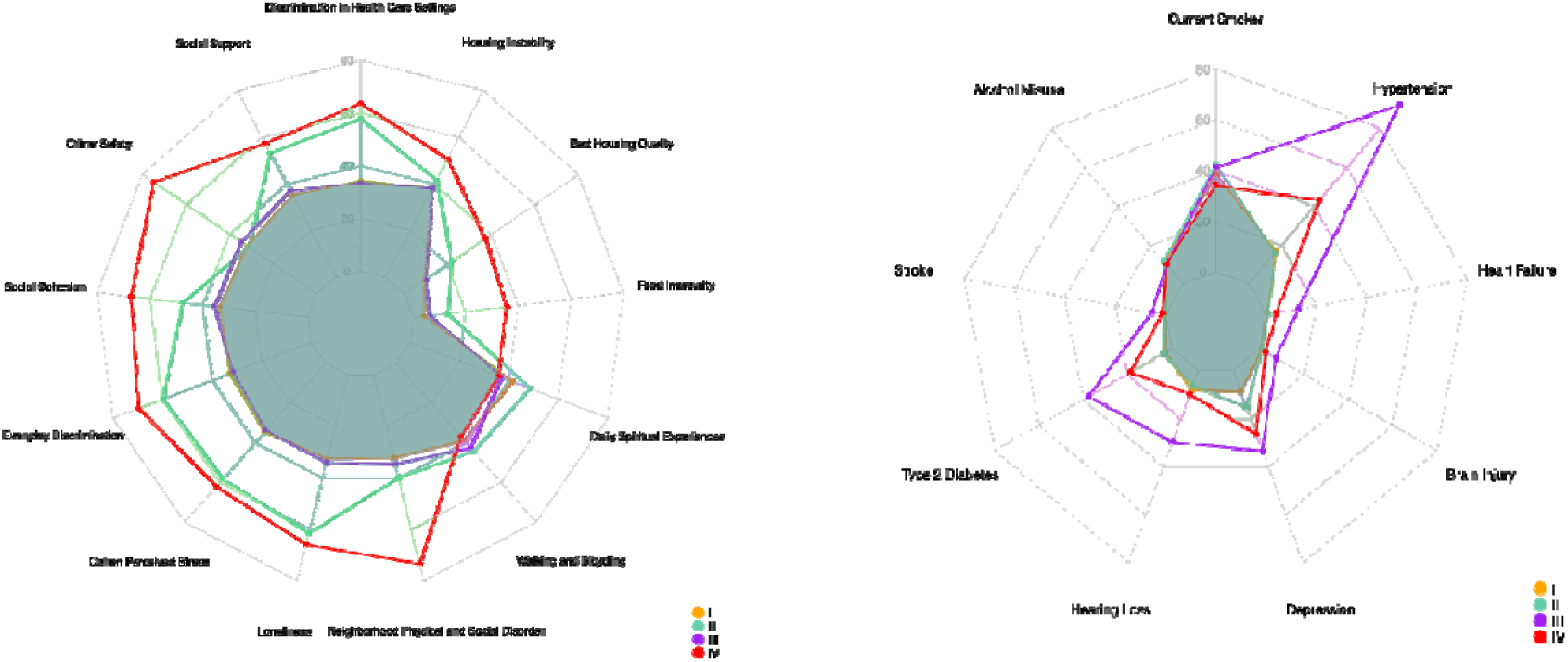
Radar plot illustrating differences in SDOH and comorbidity profiles across the four identified latent subtypes (I–IV).

**Figure 10:**
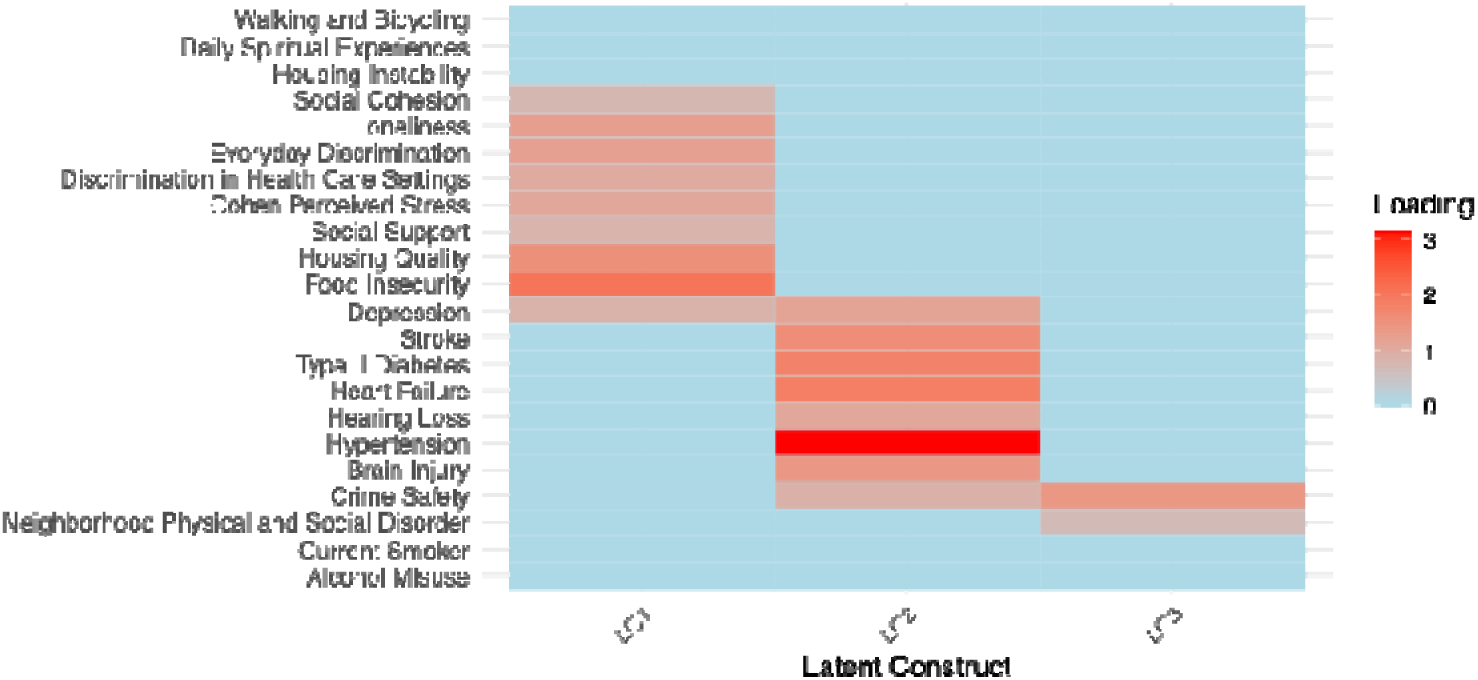
Heatmap displaying the magnitude of standardized loadings for each observed measure across the three latent constructs (LC1–LC3).

**Figure 11:**
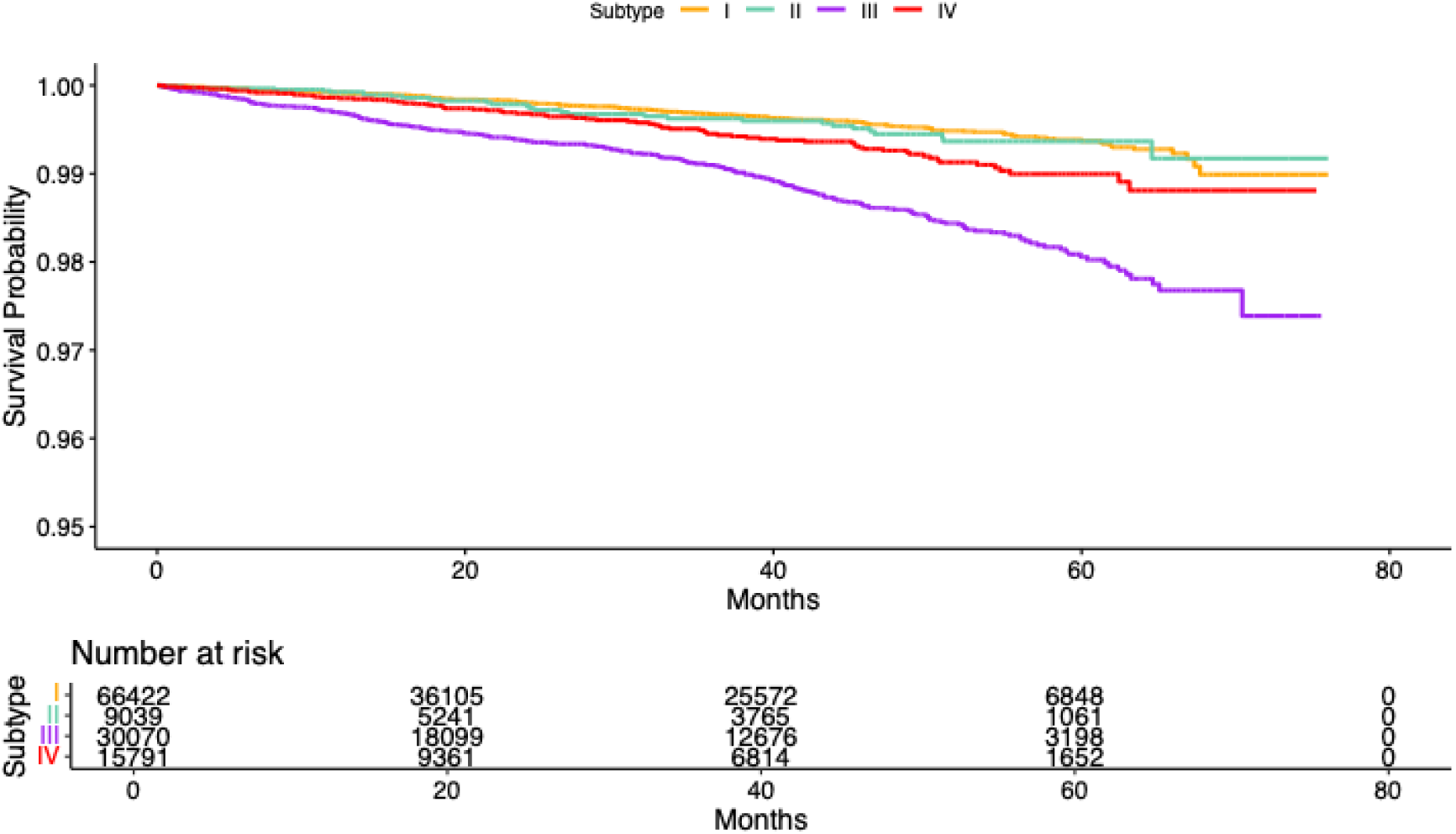
Kaplan–Meier survival curves depicting the probability of remaining free from MCI across Subtypes I–IV.

**Figure 12:**
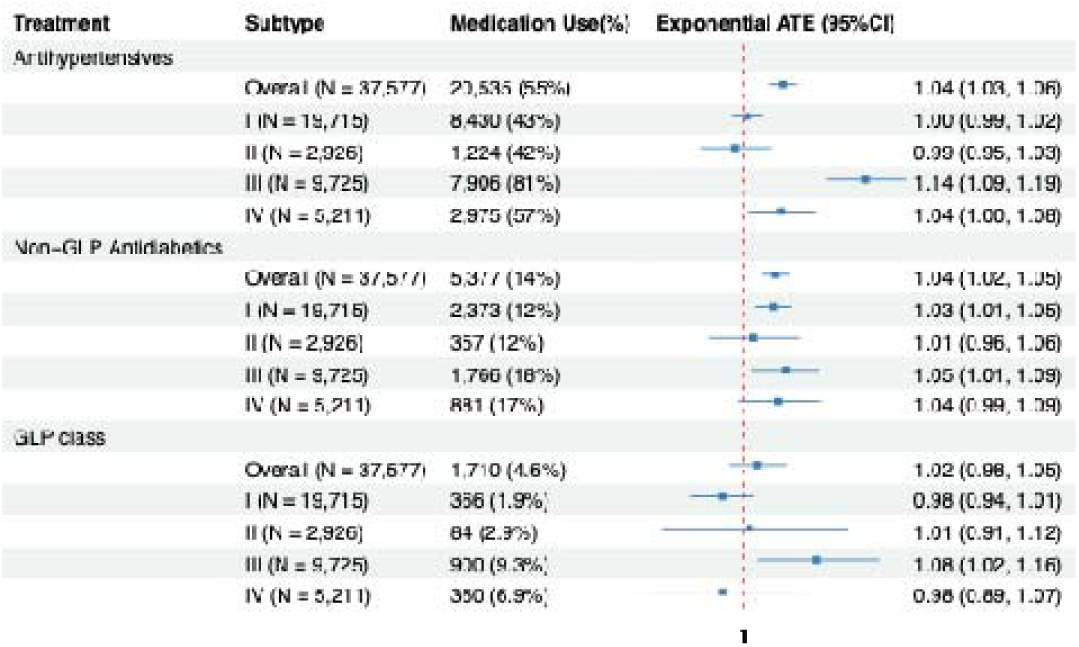
Estimated exponential average treatment effects for antihypertensive, non-GLP antidiabetic and GLP class medications across the four data-driven subtypes.

For non–GLP antidiabetic medications, the overall association was similarly modest but statistically significant (time ratio = 1.04; 95% CI, 1.02–1.05; ∼4% longer time). At the subtype level, significant associations were observed in Subtype I (time ratio = 1.03; 95% CI, 1.01–1.05; ∼3% longer time) and Subtype III (time ratio = 1.05; 95% CI, 1.01–1.09; ∼5% longer time), whereas Subtypes II and IV showed no evidence of benefit. GLP-class medications were not associated with time to MCI overall; however, Subtype III demonstrated a significant association (time ratio = 1.08; 95% CI, 1.02–1.16; ∼8% longer time). Overall, these results indicate pronounced heterogeneity in medication-associated delay of MCI onset, with Subtype III consistently exhibiting the largest estimated benefit across medication classes.

## Discussion

In this large and diverse cohort from the All of Us Research Program, we identified four data-driven subtypes of cognitively unimpaired adults using integrative modeling of comorbidities, lifestyle behaviors, and SDOH. These subtypes captured distinct multidimensional vulnerability profiles that were associated with heterogeneous risk of incident MCI and have differential estimated effect of antihypertensive and antidiabetic medication use. By integrating correlated biological, behavioral, and social risk factors into coherent latent profiles, this study provides a population-level framework for understanding heterogeneity in cognitive aging and offers insight into why cardiometabolic interventions have yielded modest and inconsistent average effects in prior dementia-prevention studies(2,10,29).

The four identified subtypes reflected distinct configurations of medical burden, behavioral risk, and social disadvantage. Subtype I represented a low-risk healthy aging profile, characterized by minimal chronic disease, low smoking and alcohol misuse, and favorable social conditions. Subtype II was marked by prominent behavioral risk and psychosocial adversity, but relatively limited cardiometabolic burden. Subtype III exhibited concentrated cardiometabolic and depressive multimorbidity, particularly hypertension, diabetes, hearing loss, and depression, with comparatively less social disadvantage. Subtype IV combined moderate medical comorbidity with pronounced social and environmental adversity, including neighborhood disorder, food insecurity, and housing instability.

These patterns align with prior epidemiologic studies of comorbidity and social disadvantage associated with adverse cognitive outcomes(8,12,30). Unlike studies that model comorbidities or SDOH as covariates or examine single domains in isolation(2,31–33), our integrative approach treats these factors as defining dimensions of heterogeneity, yielding subtypes that more closely reflect real-world co-occurring risk profiles.

Consistent with epidemiologic evidence, the subtypes showed marked differences in risk of incident MCI. Subtype III exhibited the highest hazard, aligning with evidence that cardiometabolic and vascular disease burden strongly predicts cognitive impairment and dementia(8,32,34). Subtype IV also had substantially elevated risk, supporting links between chronic social and environmental disadvantage and accelerated cognitive aging (2,33,35). In contrast, Subtype II showed a more modest increase in risk, while Subtype I maintained the most favorable MCI-free survival trajectory.

Notably, the relatively modest MCI risk observed in Subtype II, despite substantial behavioral and psychosocial adversity, may reflect delayed manifestation of stress-related effects, unmeasured intermediate conditions, or attenuation due to competing risks, differential healthcare engagement, or under-ascertainment of cognitive diagnoses(12,30,35).

We observed substantial heterogeneity in the estimated effect of cardiometabolic medication use. Subtype III consistently exhibited the largest estimated effect by antihypertensive, non–GLP antidiabetic, and GLP-class medications, suggesting that pharmacologic modification of cardiometabolic risk factors may be most impactful among individuals with substantial underlying comorbidity burden. This pattern is concordant with randomized evidence demonstrating that intensive blood pressure control reduces incident MCI among individuals at elevated vascular risk(14) and metformin improves memory performance(36). In contrast, Subtypes II and IV showed limited or selective effects by pharmacologic therapy, suggesting that when cognitive risk is driven primarily by behavioral or structural adversity, medication alone may be insufficient to meaningfully alter trajectories. Subtype I showed modest estimated effects by non–GLP antidiabetic medications despite low baseline MCI risk, raising the possibility that some cardiometabolic therapies may influence cognitive aging through mechanisms beyond prevention of vascular events, such as effects on insulin signaling, inflammation, or cerebral metabolism(21,37). This interpretation aligns with metformin clinical trials in cognitive aging, including Metformin in Mild Cognitive Impairment and TAME, which target insulin resistance and inflammation as upstream mechanisms of neurodegeneration (36,38).

Importantly, these findings should be interpreted as estimated causal effects carefully. Although we applied a double/debiased machine learning causal framework with inverse probability of censoring weights to mitigate confounding and selection bias, residual confounding related to disease severity, medication indication, adherence, healthcare access, and treatment duration cannot be excluded. Failure to account for underlying vulnerability profiles may contribute to the modest and inconsistent average effects reported in prior observational and interventional studies(10,21).

The latent constructs identified by the integrative model highlight distinct yet interacting pathways linking subtype profiles to cognitive risk and inform targeted prevention strategies. The construct dominated by cardiometabolic and depressive multimorbidity likely reflects vascular, metabolic, and inflammatory mechanisms contributing to neurodegeneration(34), suggesting that individuals in Subtype III may benefit most from intensified cardiometabolic and neurological disease management. In contrast, the construct characterized by psychosocial stress, discrimination, loneliness, and material hardship may act through chronic stress–related pathways, including hypothalamic–pituitary–adrenal axis dysregulation and reduced cognitive reserve(2), indicating that Subtype II may respond best to psychosocial and community-based interventions. The construct reflecting neighborhood disorder and safety concerns likely affects cognition indirectly by limiting physical activity, social engagement, and access to health-promoting resources(35), underscoring the need for structural interventions in Subtype IV. These pathways likely interact over the life course, reinforcing the value of multidimensional risk stratification for mechanism-informed and targeted dementia prevention.

Together, these findings underscore that dementia prevention strategies should be aligned with dominant vulnerability pathways rather than applied uniformly across heterogeneous populations. Integrative subtyping frameworks embedded within large-scale health systems can identify individuals most likely to benefit from pharmacologic modification of upstream cardiometabolic risk factors, while distinguishing subgroups for whom behavioral or structural interventions addressing psychosocial and environmental risk may be more relevant. By explicitly modeling heterogeneity in both baseline vulnerability and treatment responsiveness, this approach supports subgroup-enriched pragmatic trials and advances a more mechanism-informed, targeted, and equitable framework for precision prevention in cognitive aging.

Importantly, this population-level perspective is complementary to biomarker-based approaches. Whereas biomarkers capture downstream, disease-specific biological processes, integrative subtyping focuses on upstream vulnerability relevant to early risk stratification and prevention, together providing a more comprehensive foundation for dementia prevention across the life course(2).

Several limitations warrant consideration. First, the analysis focused on all-cause MCI, and information on etiologic MCI subtypes and severity was unavailable, as standardized cognitive test scores or continuous measures of cognitive performance were not incorporated. Second, cognitive, neuroimaging, and molecular biomarkers were not included; accordingly, the identified subtypes should be interpreted as population-level vulnerability profiles rather than direct proxies for underlying neuropathology(7). Third, medication exposure was defined using baseline prescription records and did not capture dose, adherence, or changes over follow-up. Fourth, subtypes were derived from baseline measures and therefore do not capture longitudinal changes that may influence MCI risk or treatment responsiveness. Also, although the All of Us cohort enhances generalizability through its size and diversity(15), healthy volunteer bias, differential healthcare access, and diagnostic ascertainment may affect absolute risk estimates. Last, subtype definitions are model-dependent, and alternative modeling choices or the inclusion of additional modalities could yield different subgroup structures. Independent validation in external cohorts such as the UK Biobank is therefore needed to assess the robustness, reproducibility, and generalizability of the identified subtypes.

## Conclusion

In summary, this study demonstrates that integrative, data-driven subtyping based on comorbidities, lifestyle behaviors, and social determinants of health can uncover clinically and socially meaningful heterogeneity in cognitive aging trajectories. The identified subtypes differed substantially in risk of incident MCI and in estimated associations with cardiometabolic therapies, highlighting the importance of aligning prevention strategies with dominant vulnerability pathways. By explicitly recognizing that cognitive risk emerges from interacting biological mechanisms, modifiable health behaviors, and structural determinants, this multidimensional framework provides a foundation for more targeted, equitable, and effective approaches to dementia prevention.

## Data Availability

All data produced are available online at AllofUs Research Program

## List of abbreviations

ATE: Average Treatment Effect
AUDIT-C: Alcohol Use Disorders Identification Test–Consumption
CI: Confidence Interval
CU: Cognitively Unimpaired
DoubleML: Double/Debiased Machine Learning
EHR: Electronic Health Record
GATE: Group Average Treatment Effect
GLP: Glucagon-Like Peptide
HR: Hazard Ratio
IC: Information Criterion
IRM: Interactive Regression Model
LC: Latent Construct
MCI: Mild Cognitive Impairment
MCMC: Markov Chain Monte Carlo
MINDS: Mixed Integrative Data Subtyping
OMOP: Observational Medical Outcomes Partnership
PANES: Physical Activity Neighborhood Environment Scale
SDOH: Social Determinants of Health
SNOMED CT: Systematized Nomenclature of Medicine Clinical Terms

## Declarations

### Ethics approval and consent to participate

Non applicable.

### Consent for publication

Non applicable.

### Availability of data and materials

The data that support the findings of this study are available from the National Institutes of Health’s All of Us Research Program upon reasonable request and with permission of the All of Us Research Program.

### Authors’ contributions

YZ conducted the analyses and drafted the manuscript. KM contributed to clinical interpretation and critical revisions. YW conceived and supervised the study and revised the manuscript. All authors approved the final version.

## Acknowledgements

We gratefully acknowledge All of Us participants for their contributions, without whom this research would not have been possible. We also thank the National Institutes of Health’s All of Us Research Program for making available the participant data examined in this study.

